# No difference in risk of hospitalisation between reported cases of the SARS-CoV-2 Delta variant and Alpha variant in Norway

**DOI:** 10.1101/2021.09.02.21263014

**Authors:** Lamprini Veneti, Beatriz Valcarcel Salamanca, Elina Seppälä, Jostein Starrfelt, Margrethe Larsdatter Storm, Karoline Bragstad, Olav Hungnes, Håkon Bøås, Reidar Kvåle, Line Vold, Karin Nygård, Eirik Alnes Buanes, Robert Whittaker

**Author notes:** Corresponding author: Robert Whittaker, Norwegian Institute of Public Health, Lovisenberggata 8, 0456, Oslo, Norway.

## Abstract

**Objectives:** To estimate the risk of hospitalisation among reported cases of the Delta-variant of SARS-CoV-2 compared to the Alpha variant in Norway. We also estimated the risk of hospitalisation by vaccination status.

**Methods:** We conducted a cohort study on laboratory-confirmed cases of SARS-CoV-2 in Norway, diagnosed between 3 May and 15 August 2021. We calculated adjusted risk ratios (aRR) with 95% confidence intervals (CIs) using multivariable binomial regression, accounting for variant, vaccination status, demographic characteristics, week of sampling and underlying comorbidities.

**Results:** We included 7,977 cases of Delta and 12,078 cases of Alpha. Overall, 347 (1.7%) cases were hospitalised. The aRR of hospitalisation for Delta compared to Alpha was 0.97 (95%CI 0.76–1.23). Partially vaccinated cases had a 72% reduced risk of hospitalisation (95%CI 59%–82%), and fully vaccinated cases had a 76% reduced risk (95%CI 61%–85%), compared to unvaccinated cases.

**Conclusions:** We found no difference in the risk of hospitalisation for Delta cases compared to Alpha cases in Norway. Further research from a wide variety of settings is needed to better understand the association between the Delta variant and severe disease. Our results support the notion that partially and fully vaccinated persons are highly protected against hospitalisation with COVID-19.

**Highlights:**

- The SARS-CoV-2 Delta variant has dominated in Norway since July 2021
- There was no difference in the risk of hospitalisation for Delta cases compared to Alpha
- Partially and fully vaccinated cases had >70% decreased risk of hospitalisation

## Background

Multiple variants of severe acute respiratory syndrome coronavirus 2 (SARS-CoV-2), the causative agent of COVID-19, have been observed worldwide. Some of these variants have been designated as variants of concern (VOC), defined by the WHO as variants associated with increased transmissibility, increased disease severity or change in clinical disease presentation, and/or decreased effectiveness of public health and social measures or available diagnostics, vaccines, and therapeutics [1]. Variants of concern include the Alpha variant (Phylogenetic Assignment of Named Global Outbreak (Pango) lineage designation B.1.1.7; earliest documented samples from the United Kingdom (UK) in September 2020), and the Delta variant (Pango lineage B.1.617.2; earliest documented samples from India in October 2020) [1]. Since their emergence, both variants have spread worldwide [2].

In Norway (population 5.4 million), testing activity for COVID-19 is high, with consistently 3–5% of the population tested weekly (defined as one or more tests per person within a seven-day period) since March 2021. Mathematical modelling estimates that consistently over 50% of all cases weekly have been detected since late 2020 [3]. Sequencing capacity in Norwegian laboratories was rapidly scaled up from early December 2020, and the capacity to screen for variants or perform whole genome sequencing (WGS) was further increased following reports of widespread transmission of the Alpha variant in the UK. Since early April 2021, over 70% of cases diagnosed have available data on the variant of SARS-CoV-2 that caused their infection.

Alpha has been shown to be more easily transmitted than non-VOC variants [4], and was the dominant circulating SARS-CoV-2 variant in Norway during the third wave of infections in the winter and spring of 2021. It was also associated with a 1.9-fold increased risk of hospitalisation compared to non-VOC variants [5]. Similar associations were observed in other European countries [6, 7]. Local and national non-pharmaceutical interventions and increasing vaccination coverage gradually decreased transmission, and Norway started its national reopening plan during the spring [3].

The first case of the Delta variant was diagnosed in Norway in April 2021, and local transmission was first evident in the beginning of May. Delta superseded Alpha as the dominant circulating variant in early July, accounting for over 90% of new infections by the end of that month. This coincided with the start of the fourth wave of SARS-CoV-2 infections, and a subsequent increase in the number of new hospitalisations [3]. There is evidence of increased transmissibility [8-10] and lower vaccine effectiveness against infection [11-13] for Delta compared to Alpha. In addition, studies from Scotland [12], England [14] and Ontario, Canada [15] have suggested that infection with the Delta variant increased the risk of hospitalisation by 1.5 to approximately 2-fold in those settings.

In order to understand the impact of the Delta variant on the burden of COVID-19 in Norway, and support preparedness planning in the hospital sector, we used linked individual-level data to estimate the risk of hospitalisation among reported cases of the Delta variant compared to reported cases of the Alpha variant, accounting for demographic characteristics, vaccination status and underlying comorbidities. We also estimated the risk of hospitalisation by vaccination status.

## Methods

### Data sources and study design

We obtained data from the Norwegian national preparedness registry for COVID-19 [16]. The preparedness registry contains individual-level data from central health registries, national clinical registries and other national administrative registries. It covers all residents in Norway, and includes data on all laboratory-confirmed cases of COVID-19 in Norway, all hospitalisations among cases and COVID-19 vaccinations (supplementary materials, part 1).

We conducted a cohort study, including cases who tested positive for SARS-CoV-2 between 3 May (week 18) and 15 August (week 32) 2021, who had a national identity number registered, and who had been infected with the Alpha or Delta variant, confirmed by PCR screening assays or whole genome sequencing (WGS). The laboratory testing for variants of SARS-CoV-2 in Norway has been described in detail⍰telsewhere [5]. Variants other than Delta and Alpha were infrequently detected in Norway during the study period [3]. We extracted data up to 30 August 2021, ensuring at least 15 days of follow-up since last date of sampling.

### Definitions

#### Hospitalisation

We defined hospitalisation as hospital admission following a positive SARS-CoV-2 test, where COVID-19 was reported as the main cause of admission. Cases hospitalised with other or unknown main cause of admission were excluded from the study population in order to avoid bias. All admissions to hospital, regardless of the length of stay, were included.

#### Vaccination status

In Norway, Comirnaty (BioNTech-Pfizer, Mainz, Germany/New York, United States) and Spikevax (mRNA-1273, Moderna, Cambridge, United States) are the two most frequently administered vaccines. Vaxzevria (AstraZeneca, Cambridge, United Kingdom) was initially included in the vaccine programme, but removed in March 2021. The Janssen vaccine (Janssen Vaccines, Leiden, Netherlands) is not part of the national vaccination programme in Norway, and is only given under certain circumstances [17]. We defined SARS-CoV-2 cases according to their vaccination status:

1. Those unvaccinated with a COVID-19 vaccine before positive test.
2. Those vaccinated with 1 dose of a COVID-19 vaccine less than 21 days before positive test.
3. Partially vaccinated – those who tested positive at least 21 days after their first dose of a COVID-19 vaccine, and less than 7 days after the second dose.
4. Fully vaccinated – those who tested positive at least 7 days after their second dose with at least the recommended minimum interval between doses depending on the type of vaccine [17], or 7 days after their first dose if they had previously been diagnosed with a SARS-CoV-2 infection at least 21 days before vaccination. Cases who received the Janssen vaccine were considered fully vaccinated 21 days after one dose.

#### Underlying comorbidities with increased risk of severe COVID-19

Some people have underlying comorbidities that cause them to have a moderate or high risk of severe COVID-19, regardless of age. These individuals were prioritised for vaccination [18]. We categorised cases into three groups: i) no underlying comorbidities, ii) medium risk comorbidity and iii) high risk comorbidity, as detailed in supplementary materials, part 1.

### Data analysis

We described cases in terms of variants, vaccination status, demographic characteristics, underlying comorbidities and hospitalisation. We also described cases in terms of admission to an intensive care unit and COVID-19 related deaths [19].

### Statistical analysis

We calculated adjusted risk ratios (aRR) with 95% confidence intervals (CIs) using multivariable binomial regression. Variables considered in our analysis were variant (Alpha or Delta), vaccination status (4 levels), age (4 age groups), sex, country of birth (3 levels), period of sampling (biweekly as categorical variable, and week as continuous variable), county of residence (12 levels), and underlying comorbidities (3 levels). Model selection for the multivariable binomial regression was conducted using the likelihood ratio test and the Akaike Information Criterion. We kept the variables variant (due to the main aim of the study) and sex (main demographic characteristic) in our multivariable analysis even if they were not significant. We also checked for interactions between our co-variates by including interaction terms in our models. We conducted the main analysis separately for some of the groups of variables (stratified analysis) to ensure that the associations remained robust.

In addition to our main analysis, we conducted a number of sensitivity analyses by extending or restricting our study population (for example, including only cases who had WGS results), by adjusting our outcome definitions (for example, including all cases who were hospitalised regardless of main cause of admission) and by changing our analysis method (for example, using Cox regression) to further explore if our main results were robust (supplementary materials, part 2.1).

We also assessed the power of our study to detect a range of potential effect sizes for the risk of hospitalisation with the Delta variant, compared to Alpha (supplementary materials, part 2.2).

Statistical analysis was performed in Stata version 16 (Stata Corporation, College Station, Texas, US), and R version 4.1.0.

### Ethics

Ethical approval for this study was granted by Regional Committees for Medical Research Ethics - South East Norway, reference number 249509. The need for informed consent was waived by the ethics committee.

## Results

### Description of cohort

There were 30,386 cases of COVID-19 diagnosed between 3 May and 15 August 2021 in Norway. Of these we excluded 644 cases who did not have a national identity number, as well as 111 hospitalised with other main cause of admission than COVID-19 and three with unknown main cause of hospital admission. Of the remaining 29,628, 21,691 (73%) had data on virus variant. The proportion of cases with data on virus variant varied from 64% to 80% per week during the study period. Delta superseded Alpha as the predominant circulating variant in week 27 (Fig 1). By August only sporadic cases of Alpha-variant were detected. In supplementary materials, part 2.3 we present characteristics of cases with data on virus variant compared to all notified cases.

**Figure 1.**
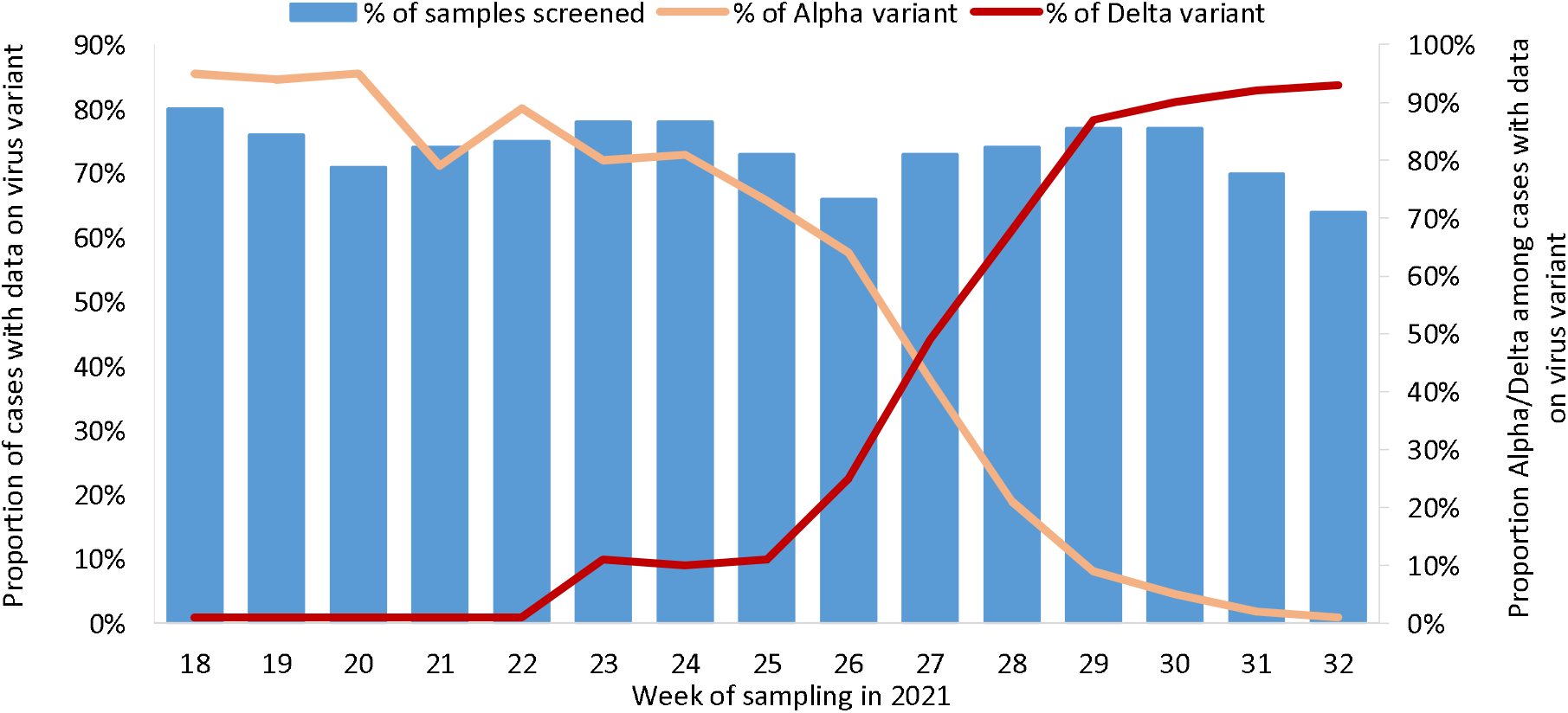
Proportion of SARS-CoV-2 cases with data on virus variant, and proportion with Alpha and Delta, by week of sampling, Norway, 3 May – 15 August 2021.

Of the 21,691 cases, 7,977 (37%) were Delta and 12,078 (56%) Alpha. There were also six cases of the Beta variant (B.1.351), four of the Gamma variant (P.1) and 384 non-VOC, while 1,242 could not clearly be categorised as one of the VOC or a non-VOC based on the available information. The 20,055 Delta and Alpha cases are henceforth referred to as our study cohort. Characteristics of the study cohort are presented in Table 1. The median age was 26 years (interquartile range (IQR): 18– 37) for Delta cases and 24 years (IQR: 18–39) for Alpha cases. WGS was used to determine the variant in 3,040 (38%) Delta cases, and 6,557 (54%) Alpha cases. At the time of diagnosis, 15,140 cases (75%) were not vaccinated, 1,722 (9%) were vaccinated with 1 dose less than 21 days before positive test, 2,386 (12%) were partially vaccinated and 807 (4%) were fully vaccinated. More details on vaccine types among partially and fully vaccinated cases by variant is presented in supplementary materials, part 2.4.

**Table 1.** Distribution of SARS-CoV-2 cases in study cohort by detected variants for different characteristics and proportion hospitalised, Norway, 3 May – 15 August 2021

| Characteristics |  | Study cohort | Variant type (% by characteristic) |  | Hospitalised cases (% of diagnosed cases) |  |  |
| --- | --- | --- | --- | --- | --- | --- | --- |
|  |  |  | Alpha | Delta | Alpha | Delta | Total |
| **Total** |  | 20,055 | 12,078 (60%) | 7,977 (40%) | 240 (2.0%) | 107 (1.3%) | 347 (1.7%) |
| **Sex** | **Female** | 9,433 | 5,750 (48%) | 3,683 (46%) | 104 (1.8%) | 53 (1.4%) | 157 (1.7%) |
|  | **Male** | 10,622 | 6,328 (52%) | 4,294 (54%) | 136 (2.2%) | 54 (1.3%) | 190 (1.8%) |
| **Age group** | **0-24 years** | 9,986 | 6,292 (52%) | 3,964 (46%) | 11 (0.2%) | 13 (0.4%) | 24 (0.2%) |
|  | **25-44 years** | 6,559 | 3,523 (29%) | 3,036 (38%) | 69 (2.0%) | 44 (1.5%) | 113 (1.7%) |
|  | **45-64 years** | 3,060 | 2,003 (17%) | 1,057 (13%) | 120 (6.0%) | 35 (3.3%) | 155 (5.1%) |
|  | **≥65 years** | 450 | 260 (2.2%) | 190 (2.4%) | 40 (15%) | 15 (7.9%) | 55 (12%) |
| **Norwegian born** | **Yes** | 14,106 | 8,743 (72%) | 5,363 (67%) | 131 (1.5%) | 49 (0.9%) | 180 (1.3%) |
|  | **No** | 5,802 | 3,241 (27%) | 2,561 (32%) | 101 (3.1%) | 55 (2.2%) | 156 (2.7%) |
|  | **Unknown** | 147 | 94 (0.8%) | 53 (0.7%) | 8 (8.5%) | 3 (5.7%) | 11 (7.5%) |
| **Risk for severe COVID-19\*** | **No underlying comorbidities** | 18,291 | 10,962 (91%) | 7,329 (92%) | 173 (1.6%) | 86 (1.2%) | 259 (1.4%) |
|  | **Medium risk comorbidity** | 1,637 | 1,040 (8.6%) | 597 (7.5%) | 57 (5.5%) | 16 (2.7%) | 73 (4.5%) |
|  | **High risk comorbidity** | 127 | 76 (0.6%) | 51 (0.6%) | 10 (13%) | 5 (9.8%) | 15 (12%) |
| **Vaccination status at date of positive test** | **Not vaccinated** | 15,140 | 10,713 (89%) | 4,427 (56%) | 184 (1.7%) | 73 (1.6%) | 257 (1.7%) |
|  | **Vaccinated with 1 dose <21 days before positive test** | 1,722 | 726 (6.0%) | 996 (12%) | 35 (4.8%) | 7 (0.7%) | 42 (2.4%) |
|  | **Partially vaccinated** | 2,386 | 461 (3.8%) | 1,925 (24%) | 12 (2.6%) | 14 (0.7%) | 26 (1.1%) |
|  | **Fully vaccinated** | 807 | 178 (1.5%) | 629 (7.9%) | 9 (5.1%) | 13 (2.1%) | 22 (2.7%) |

\* Risk for severe disease based on underlying comorbidities that are associated with a moderate or high risk of serious illness regardless of age. Details on the definitions of medium and high risk categories are provided in supplementary materials, part 1.

### Risk of hospitalisation

During the study period, 347 (1.7%) cases were hospitalised with COVID-19 as main cause of hospitalisation. Among Delta cases 107 (1.3%) were hospitalised, compared to 240 (2.0%) among Alpha cases (Table 1). The median time from testing to hospitalisation was slightly shorter for Delta cases (5 days, IQR: 1–7) than Alpha (6 days, IQR: 3–8.5; Wilcoxon rank-sum p value = 0.016).

In the univariate analysis, the crude RR for hospitalisation among those infected with Delta compared to Alpha was 0.68 (95%CI 0.54–0.85) suggesting a lower risk for hospitalisation among Delta cases. In our multivariable model, after adjusting for sex, age group, country of birth, vaccination status and underlying comorbidities, no difference was found in the risk of hospitalisation between Delta and Alpha, with an aRR of hospitalisation of 0.97 (95%CI 0.76–1.23) (Table 2). Week of sampling and county of residence were not significant predictors in the multivariable model and were excluded from the final model. When we checked for interactions, only an interaction between age group and vaccination status was detected. In order to simplify our main results presented here, we decided to not include the interaction term in our main model. The association between vaccination status and hospitalisation stratified by age group and other different groups is presented in supplementary materials, part 2.5. No interaction was found in our main multivariable analysis between variant and vaccination status. In Table 3, we present the stratified aRR estimates for Delta compared to Alpha which confirmed the findings in the main analysis. The aRR of hospitalisation among unvaccinated cases for Delta compared to Alpha was 1.10 (95%CI: 0.84–1.45). Our results were robust in all our sensitivity analyses (supplementary materials, part 2.1).

**Table 2.** Risk ratios for hospitalisation with COVID-19 as main cause from univariate and multivariable binomial regression adjusted for variant, sex, age group, country of birth, underlying comorbidities, and vaccination status at date of positive test, Norway, 3 May – 15 August 2021.

|  |  | Hospitalisation |  |  |  |
| --- | --- | --- | --- | --- | --- |
|  |  | No | Yes (%) | Crude risk ratio (95%CI) | Adjusted risk ratio (95%CI) |
| **Variant** | **Alpha** | 11,838 | 240 (2.0 %) | Ref | Ref |
|  | **Delta** | 7,870 | 107 (1.3 %) | 0.68 (0.54-0.85) | 0.97 (0.76-1.23) |
| **Sex** | **Female** | 9,276 | 157 (1.7%) | Ref | Ref |
|  | **Male** | 10,432 | 190 (1.8%) | 1.07 (0.87-1.33) | 1.05 (0.85-1.28) |
| **Age group** | **25-44 years** | 6,446 | 24 (0.2%) | Ref | Ref |
|  | **0-24 years** | 9,962 | 113 (1.7%) | 0.14 (0.09-0.22) | 0.14 (0.09-0.22) |
|  | **45-64 years** | 2,905 | 155 (5.1%) | 2.94 (2.32-3.73) | 3.18 (2.48-4.08) |
|  | **≥65 years** | 395 | 55 (12%) | 7.09 (5.21-9.65) | 10.27 (7.06-14.95) |
| **Norwegian born** | **Yes** | 13,926 | 180 (1.3%) | Ref | Ref |
|  | **No** | 5,646 | 156 (2.7%) | 2.11 (1.70-2.61) | 1.38 (1.12-1.70) |
|  | **Unknown** | 136 | 11 (7.5%) | 5.86 (3.26-10.54) | 1.23 (0.66-2.30) |
| **Risk for severe COVID-19 \*** | **No underlying comorbidities** | 18,032 | 259 (1.4) | Ref | Ref |
|  | **Medium risk comorbidity** | 1,564 | 73 (4.5%) | 3.15 (2.44-4.06) | 1.75 (1.33-2.30) |
|  | **High risk comorbidity** | 112 | 15 (12%) | 8.34 (5.11-13.62) | 2.94 (1.74-4.99) |
| **Vaccination status at date of positive test** | **Not vaccinated** | 14,883 | 257 (1.7%) | Ref | Ref |
|  | **Vaccinated with 1 dose <21 days before positive test** | 1,680 | 42 (2.4%) | 1.44 (1.04-1.98) | 0.78 (0.56-1.08) |
|  | **Partially vaccinated** | 2,360 | 26 (1.1%) | 0.64 (0.43-0.96) | 0.28 (0.18-0.41) |
|  | **Fully vaccinated** | 785 | 22 (2.7%) | 1.61 (1.05-2.47) | 0.24 (0.15-0.39) |

CI: confidence intervals. Week of sampling and county of residence were not significant predictors in the multivariable model and not included in the final model. \* Risk for severe disease based on underlying comorbidities that are associated with a moderate or high risk of serious illness regardless of age. Details on the definitions of medium and high risk categories are provided in supplementary materials, part 1.

**Table 3.**
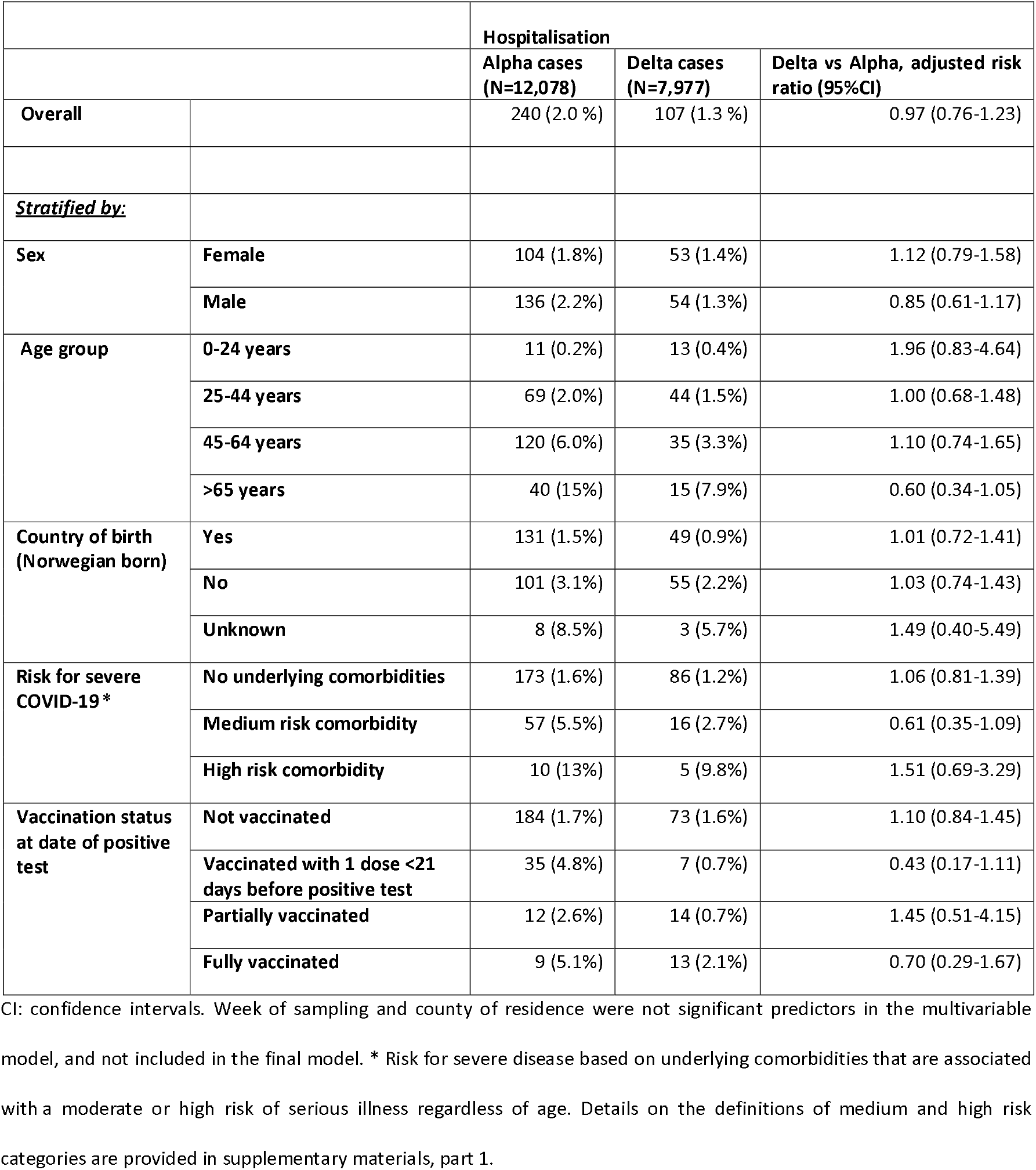
Association between hospitalisation and infection with Delta variant compared to Alpha variant of SARS-CoV-2 (multivariable binomial regression adjusted for demographic characteristics and underlying comorbidities), overall and stratified by sex, age groups, country of birth, underlying comorbidities and vaccination status at date of positive test, Norway, 3 May – 15 August 2021.

The crude RR for hospitalisation among fully vaccinated compared to unvaccinated cases changed when we adjusted for other factors due to confounding. In our multivariable model, after adjusting for sex, age group, country of birth, variant and underlying comorbidities, partially vaccinated cases had a 72% reduced risk of hospitalisation (95%CI 59%–82%) and fully vaccinated had a 76% reduced risk of hospitalisation (95%CI 61%–85%) compared to unvaccinated cases (Table 2). In supplementary materials, part 2.5 we present stratified estimates for the risk of hospitalisation by vaccination status.

### Admission to ICU and COVID-19 related deaths

Among the 107 individuals hospitalised with Delta, 16 (15%) of were admitted to ICU compared to 40 (17%) of 240 individuals hospitalised with Alpha. Among the 56 cases admitted to ICU, 40 were unvaccinated and 14 had been vaccinated <21 days before positive test. There were five COVID-19 related deaths reported among Delta cases, and 15 reported among Alpha cases. Additional analyses were not done on these outcomes due to small numbers.

## Discussion

In this study, we have analysed individual-level data on all laboratory-confirmed cases of COVID-19 in Norway and hospitalisations among cases within the study period, as well as demographic characteristics, vaccination status and underlying comorbidities. Our findings indicate no difference in the risk of hospitalisation for SARS-CoV-2 cases infected with the Delta variant compared to the Alpha variant in Norway. This contrasts with previously published estimates. An analysis from Scotland suggested an adjusted hazard ratio for hospitalisation of 1.85 (95%CI 1.39–2.47) for Delta compared to Alpha [12]. In England, a similar association was observed (hazard ratio 2.26, 95%CI 1.32–3.89) [14]. The study from England also suggested increased risk of hospitalisation in an unvaccinated cohort (hazard ratio 2.32, 95%CI 1.29–4.16), which our study also does not support. In addition, a preprint study from Ontario, Canada estimated an adjusted odds ratio for hospitalisation of 1.49 (95%CI 1.41–1.58) for Delta compared to other VOC (Alpha, Beta or Gamma) [15].

In comparing the estimates, differences between the studies must be considered. For example, the study in Ontario was based on cases diagnosed from February to June 2021, England on cases from March to May, Scotland on cases from May and our study on cases from May to August. The different settings in which these studies were conducted may have contributed to the associations observed, and warrant further study. Subtle differences in the outcome definitions between the studies also need to be considered, as well as analysis methods. However, when we ran Cox regression models, more closely reflecting the analysis by Scotland and England, our results were robust (supplementary materials, part 2.1). Other differences in analysis methods between the studies also exist. For example, Scotland adjusted for the level of socioeconomic deprivation, which was not available in our study, although this was not a significant predictor of hospitalisation in the model from Scotland [12]. Conversely, we adjusted for country of birth, which has been associated with increased risk of hospitalisation in Norway [20]. The study from England additionally adjusted for index of multiple deprivation and international traveller status, but not directly for underlying comorbidities [14].

Our results highlight the importance of taking local epidemiological characteristics into account, when endeavouring to understand the effect that different variants have on the COVID-19 epidemic in different settings. Our results are representative of a young cohort of SARS-CoV-2 cases in a country in which the health system operated well within capacity, and where there was high vaccination coverage among populations at greater risk of severe COVID-19. Vaccination coverage was also steadily increasing as Delta superseded Alfa as the dominant variant [3]. Also, the variation in the estimates presented in different studies emphasises the need for more research from a wide variety of settings to better understand the association between the Delta variant and severe disease.

In this study we did not have access to data on clinical disease severity among cases, and the number of ICU admissions and COVID-19 related deaths were low in both groups. Our results therefore cannot directly conclude as to whether there is a difference in virulence for Delta compared to Alpha. What our results do suggest is that other factors, such as age, country of birth, underlying comorbidities and vaccination status continue to dictate the risk of hospitalisation among SARS-CoV-2 cases in Norway. However, even if the risk of hospitalisation among Delta and Alpha cases in Norway is similar, risk of infection with Delta is higher given evidence of increased transmissibility [8-10] and lower vaccine effectiveness against infection [11-13]. This must be considered as prevention and control measures are weighed up in view of the burden of disease in society, capacity in the health care system and progress of vaccination programmes. In Norway, vaccine effectiveness against laboratory confirmed infection with the Delta variant has been estimated to be 22% among partially vaccinated and 65% among fully vaccinated persons [13]. Our results suggest that partially and fully vaccinated cases infected with the Delta variant are highly protected against hospitalisation, in line with published estimates from elsewhere [12, 21, 22]. This highlights the importance of ensuring high vaccination uptake. As of the end of August 2021, first dose vaccination coverage in Norway among persons 18 years and older is 89%, while two dose coverage is 70% [3]. The main vaccines administered during the study period were the mRNA vaccines Comirnaty and Spikevax. The vast majority of vaccinated cases received Comirnaty, which did not allow us to investigate whether the type of vaccine had an additional impact on the risk of hospitalisation (supplementary materials, part 2.4).

Our study has several strengths. Firstly, testing activity during the study period was high, and over 70% of all diagnosed cases had data on the variant of SARS-CoV-2 that they were infected with. Also, during the study period, hospitals in Norway functioned within capacity, criteria for hospital admission were consistent and hospital treatment was available to all those who would benefit. In addition, aside from local restrictions in the event of outbreaks, there were no notable lockdowns during the study period. Our results were also robust when we conducted various sensitivity analyses, such as restricting our study population to only cases sequenced with WGS.

A potential source of bias is if there are systematic differences related to the variants among diagnosed and non-diagnosed cases. For example, if a larger proportion of milder cases were diagnosed in our Delta cohort, this could underestimate the risk of hospitalisation for Delta compared to Alpha. As we did not have data on relevant parameters that would have helped us to explore this further, such as clinical disease severity or viral loads, we cannot rule out this bias. However, such bias should be considered in light of the consistent COVID-19 testing strategy in Norway during the study period, with tests available free of charge for everyone, including those with mild or no symptoms, close contacts, and individuals in quarantine. In addition, it has been suggested that, when comparing two variants for a post-infection outcome at a time when one variant is in the process of supplanting the other, that in fact a relatively larger proportion of severe cases of the new variant (in this case Delta) could be diagnosed [23].

There are also some limitations with our analysis. While our sample size was marginally larger than the study from Scotland [12], both in terms of number of cases overall and number of Delta cases, our power calculations indicated that our study may be underpowered if Delta was associated with a small increased risk of hospitalisation compared to Alpha (supplementary materials, part 2.2). However, given our estimated aRR and 95%CI, this seems unlikely in our study setting. In addition, the method used to determine underlying comorbidities will likely underestimate the true prevalence, as only individuals that have been in contact with health services are identified. Data on medications used and procedure codes are currently not taken into account, which would improve the definitions and detect more individuals with underlying comorbidities. Finally, it should be noted that the reported main cause of hospitalisation is a clinical assessment. We cannot rule out that COVID-19 may have been a contributing factor to admission for some patients reported as having another main cause of hospitalisation. However, there is no reason to believe that the latter two limitations would differ between patients infected with different variants. We also conducted a sensitivity analysis involving all cases hospitalised regardless of main cause, and our results were robust.

Our findings indicate no difference in the risk of hospitalisation for cases infected with the Delta variant of SARS-CoV-2 compared to Alpha in Norway. This is a more encouraging finding for ongoing preparedness planning in hospitals than previous studies, although more research from a wide variety of settings is needed to further understand the association between the Delta variant and severe disease. Data on protection against severe disease are crucial to guide future vaccination strategy, and the results from this study and others indicate that partially and fully vaccinated persons are highly protected against hospitalisation with COVID-19.

## Data Availability

The datasets analysed during the current study come from the national emergency preparedness registry for COVID-19, housed at the Norwegian Institute of Public Health. The preparedness registry comprises data from a variety of central health registries, national clinical registries and other national administrative registries. Further information on the preparedness registry, including access to data from each data source, is available at https://www.fhi.no/en/id/infectious-diseases/coronavirus/emergency-preparedness-register-for-covid-19/.

## Notes and acknowledgements

### Authors’ contributions

All co-authors were involved in the conceptualisation of the study. RW drafted the study protocol and coordinated the study. MLS, EAB, KB, OH and RK contributed directly to the acquisition of data. LaVe, BVS, ES, JS, MLS, KB, OH, HB and RW contributed to data cleaning, verification and preparation. LaVe, BVS, ES, JS, HB and RW had access to the final linked dataset. LaVe conducted the statistical analysis with support from BVS, JS and RW. BVS conducted the power calculation. All co-authors contributed to the interpretation of the results. LaVe and RW drafted the manuscript. All co-authors contributed to the revision of the manuscript and approved the final version for submission.

## Acknowledgements

First and foremost, we wish to thank all those who have helped report data to the national emergency preparedness registry at the Norwegian Institute of Public Health (NIPH) throughout the pandemic. We also highly acknowledge the efforts that regional laboratories have put into establishing a routine variant screening procedure or whole genome sequencing at short notice and registration of all analysis in national registries for surveillance. Thanks also to the staff at the Virology and Bacteriology departments at NIPH involved in national variant identification and whole genome analysis of SARS-CoV-2 viruses. We also highly acknowledge the efforts of staff at hospitals around Norway to ensure the reporting of timely and complete data to the Norwegian Intensive Care and Pandemic Registry, as well as colleagues at the register itself. We would also like to thank Anja Elsrud Schou Lindman, project director for the national preparedness registry, and all those who have enabled data transfer to this registry, especially Gutorm Høgåsen at the NIPH, who has been in charge of the establishment and administration of the registry. We would also like to thank Hanne Gulseth and ‘Team risk group’ at the NIPH, who developed the data cleaning procedure for underlying comorbidities in the preparedness registry, as well as Trude Marie Lyngstad, Anders Skyrud Danielsen, Nora Dotterud and Evy Dvergsdal at the NIPH for their assistance in cleaning the data from different registries.

