## Supplementary materials for "No difference in risk of hospitalisation between reported cases of the SARS-CoV-2 Delta variant and Alpha variant in Norway"

### 1. Additional information on the data sources

The national emergency preparedness register, Beredt C19, contains individual-level data from central health registries, national clinical registries and other national administrative registries.

We included data on notified cases of laboratory-confirmed SARS-CoV-2 infection from the Norwegian Surveillance System for Communicable Diseases (MSIS). Data on virus variants came from the MSIS laboratory database (national laboratory database), which receives SARS-CoV-2 test results from all Norwegian microbiology laboratories.

We obtained data on hospitalisation following a positive SARS-CoV-2 test from the Norwegian Intensive Care and Pandemic Registry (NIPaR). All Norwegian hospitals report to NIPaR, and reporting is mandatory. For patients who contracted SARS-CoV-2 while admitted to hospital, the time of admission is set to the date of symptom onset, or date of sampling if the patient is asymptomatic. The reported main cause of hospitalisation is a clinical assessment. For patients reported with a different main cause than COVID-19, we cannot rule out that COVID-19 may have been a contributing factor for admission. Full details on the registration of hospitalised patients are available here (in Norwegian): https://helse-bergen.no/norsk-pandemiregister/registrering-i-norsk-pandemiregister-informasjon-til-ansatte.

Data on COVID-19 vaccinations came from the Norwegian Immunisation Registry, SYSVAK. Data on persons with a national identity number was drawn from the national population registry. The national identity number was essential to link data from all registries used in the analysis.

Data on underlying comorbidities, as stipulated by the national COVID-19 vaccination programme, was based on ICD-10 codes from the Norwegian Patient Registry, and ICPC-2 codes from the Norway Control and Payment of Health Reimbursement database (Table S1). The underlying comorbidities that have been defined as increasing the risk of severe COVID-19 are divided into two groups. Medium risk includes people with diseases/conditions that entail a moderate risk of severe COVID-19. This includes chronic liver disease or significant hepatic impairment, immunosuppressive therapy as in autoimmune diseases, diabetes, chronic lung disease including cystic fibrosis and severe asthma which have required the use of high dose inhaled or oral steroids within the past year, obesity with a body mass index (BMI) of ≥35 kg/m2, dementia, chronic heart and vascular disease (with the exception of high blood pressure) and stroke. High risk includes people with diseases/conditions that carry a high risk of severe COVID-19, also in younger individuals. These comorbidities include having received an organ transplant, immunodeficiency, hematological cancer in the last five years, other active cancers, ongoing or recently discontinued treatment for cancer (especially immunosuppressive therapy, radiation therapy to the lungs or cytotoxic drugs), neurological or neuromuscular diseases that cause impaired cough or lung function (e.g., ALS and cerebral palsy), Down syndrome and chronic kidney disease, or significant renal impairment.

**Table S1. ICD-10 codes from the Norwegian Patient Registry and ICPC-2 codes from the Norway Control and Payment of Health Reimbursement database used to identify cases with underlying comorbidities.**

| Underlying comorbidity | Specifications | ICD10-codes | ICPC-2 codes |
|---|---|---|---|
| Cardiovascular diseases, not including hypertension | | I05, I06, I07, I08, I09, I2, I31, I32, I34, I35, I36, I37, I39, I40, I41, I42, I43, I46, I48, I49, I50, I60, I61, I62, I63, I64, I69.1, I69.2, I69.3, I69.4, I69.8, I69.0 | K74, K75, K76, K77, K78, K82, K83, K90, K91 |

| | | | |
|---|---|---|---|
| Chronic pulmonary diseases, including asthma | | J41, J42, J43, J44, J45, J46, J47, J84, J98, E84 | R95, R96 |
| Compromised immune function | Organ transplantation, immune deficiency disorders, autoimmune conditions treated with immunosuppressants | Z94.0, Z94.1, Z94.2, Z94.3, Z94.4, Z94.8, D80, D81, D82, D83, D84, G35, M05, M08, M06, M07, M09, M13, M14, K50, K51 | |
| Neurological and musculoskeletal disorders with compromised lung or cough function | | G1, G20, G21, G23, G24, G40.5, G61.0, 70, G71, G80.0, G80.2, G80.3, F72, F73, F84.0, F84.1, Q05.0, Q05.1, Q05.2, Q05.3, Q05.04, Q05.5, Q05.6 | |
| Diabetes | | E10, E11, E12, E13, E14 | T89, T90 |
| Active cancer treatment or hematological cancer | | C81, C82, C83, C84, C85, C86, C87, C88, C89, C90, C91, C92, C93, C94, C95, C96, D45, D45, D47, C0, C1, C2, C3, C4, C5, C6, C7, C80, D32, D33, D35.2, D35.3, D35.4, D42, D43, D44.2, D44.3, D44.4 | |
| Other risk groups | Dementia, chronic kidney and liver disease, obesity | N18.3, N18.4, N18.5, K70.4, K72, F00, F01, F02, F03, G30, G31, E66 | P70, T82 |

### 2. Additional data and statistical analysis

*2.1 Sensitivity analyses*

In addition to the main analysis, we also conducted a number of sensitivity analyses to further explore our results regarding the risk of hospitalisation. The estimates from our sensitivity analyses are presented in Table S2. The methodology used was the same as the one used in the main analysis (see Table 2 in the manuscript). We modified the study population, by including or excluding some of the cases in our study cohort, and definition of hospitalisation, as well as including the variables week of sampling and county of residence in the model.

We also conducted an analysis where we used Cox regression in order to allow comparisons with other studies that have used this methodology [1, 2] but also in order to investigate if taking into account the time from sampling to hospitalisation would have influenced our estimates. The results using Cox regression were very similar to the one from binomial regression indicating that our results where robust. More details about the Cox regression model are provided below.

**Table S2. Infection with Delta variant of SARS-CoV-2 and risk of hospitalisation compared to Alpha when modifying the analysis method, study population or definition of outcome, Norway, 3 May – 15 August 2021**

| Analysis | Alpha cases | Delta cases | |
|---|---|---|---|
| | Hospitalised cases (%) | Hospitalised cases (%) | Adjusted risk ratio (95%CI) |

| | | | |
|---|---|---|---|
| Main analysis (n=20,055) * | 240 (2.0%) | 107 (1.3%) | 0.97 (0.76-1.23) |
| *Changes in analysis method, or variables included in the model* | | | |
| Main analysis when using Cox regression ** (n=20,053) | 239 (2.0%) | 106 (1.3%) | 0.98 (0.77-1.26) |
| Main analysis when including week of sampling in the model (n=20,055) | 240 (2.0%) | 107 (1.3%) | 0.87 (0.58-1.33) |
| Main analysis when including week of sampling and county of residence in the model (n=20,055) | 240 (2.0%) | 107 (1.3%) | 0.89 (0.58-1.36) |
| *Changes in study population or definition of outcome* | | | |
| When extending the analysis to cases diagnosed from week 15 to week 32, 2021, since the time when the first Delta cases were detected (n=26,302) | 435 (2.4%) | 108 (1.4%) | 1.01 (0.81-1.26) |
| Excluding 10,458 cases in our study cohort that were not sequenced with WGS (only had PCR screening result) (n=9,597) | 152 (2.3%) | 45 (1.5%) | 0.83 (0.59-1.19) |
| Including as hospitalised 62 cases that had another main cause of hospitalisation than COVID-19 (n=20,117) | 278 (2.3%) | 131 (1.6%) | 0.98 (0.78-1.21) |
| Including as non-hospitalised 62 cases that had another main cause of hospitalisation than COVID-19 (n=20,117) | 240 (2.0%) | 107 (1.3%) | 0.97 (0.76-1.23) |
| Including as hospitalised those that were hospitalised with COVID-19 (regardless of main cause) no more than 2 days before sampling or 14 days after sampling (n=20,113) | 274 (2.3%) | 131 (1.6%) | 0.99 (0.80-1.24) |
| Excluding 42 cases that had duration of hospitalisation ≤1 day (n=20,013) | 219 (1.8%) | 86 (1.1%) | 0.89 (0.69-1.16) |

WGS: whole genome sequencing; CI: confidence intervals. * Multivariable binomial regression adjusted for sex, age group, country of birth and underlying comorbidities, as in Table 2 in the manuscript. ** In this analysis we estimated adjusted hazard ratios using Cox regression

We repeated our main analysis, using Cox regression, calculating adjusted hazard ratios with 95% confidence intervals. We used the duration of time (in days) from date of positive test to hospitalisation and for non-hospitalised cases we used time from date of positive test to the end of the study period (30th of August) as time at risk. Two hospitalised cases were dropped from this analysis since they had date of hospitalisation 1-2 days before having a positive test. We also replaced the time at risk to 0.5 days for hospitalised cases who tested positive the day they were hospitalised (and therefore originally had 0 days at risk). Our estimates in Table S3 are very similar to the estimates using binomial regression.

**Table S3. Hazard ratios for hospitalisation for cases infected with the Delta variant of SARS-CoV-2 compared to Alpha using Cox regression, Norway, 3 May – 15 August 2021.**

| | | Hospitalisation | | | |
|---|---|---|---|---|---|
| | | No | Yes (%) | Crude hazard ratio (95%CI) | Adjusted hazard ratio (95%CI) |
| Variant | Alpha | 11,838 | 239 (2.0%) | Ref | Ref |

| | | | | | |
|---|---|---|---|---|---|
| | Delta | 7,870 | 106 (1.3%) | 0.97 (0.76-1.23) | 0.98 (0.77-1.26) |
| Sex | Female | 9,276 | 156 (1.7%) | Ref | Ref |
| | Male | 10,432 | 189 (1.8%) | 1.045 (0.85-1.28) | 1.03 (0.83-1.28) |
| Age group | 25-44 years | 6,446 | 113 (1.7%) | Ref | Ref |
| | 0-24 years | 9,962 | 24 (0.2%) | 0.14 (0.09-0.22) | 0.14 (0.09-0.22) |
| | 45-64 years | 2,905 | 154 (5.1%) | 3.18 (2.48-4.08) | 3.23 (2.51-4.16) |
| | ≥65 years | 395 | 54 (12%) | 10.27 (7.06-14.95) | 11.49 (7.69-17.16) |
| Norwegian born | Yes | 13,926 | 179 (1.3%) | Ref | Ref |
| | No | 5,646 | 155 (2.7%) | 1.38 (1.12-1.70) | 1.42 (1.14-1.77) |
| | Unknown | 136 | 11 (7.5%) | 1.23 (0.66-2.30) | 1.30 (0.69-2.45) |
| Risk for severe COVID-19 * | No underlying comorbidities | 18,032 | 257 (1.4) | Ref | Ref |
| | Medium risk comorbidity | 1,564 | 73 (4.5%) | 1.75 (1.33-2.30) | 1.84 (1.39-2.44) |
| | High risk comorbidity | 112 | 15 (12%) | 2.94 (1.74-4.99) | 3.48 (1.97-6.14) |
| Vaccine status at date of positive test | Not vaccinated | 14,883 | 256 (1.7%) | Ref | Ref |
| | Vaccinated with 1 dose <21 days before positive test | 1,680 | 42 (2.4%) | 0.78 (0.56-1.08) | 0.78 (0.56-1.09) |
| | Partially vaccinated | 2,360 | 25 (1.1%) | 0.28 (0.18-0.41) | 0.25 (0.16-0.38) |
| | Fully vaccinated | 785 | 22 (2.7%) | 0.24 (0.15-0.39) | 0.22 (0.13-0.36) |

* Risk for severe disease based on underlying comorbidities that are associated with a moderate or high risk of serious illness regardless of age. Details on the definitions of medium and high risk categories are provided in supplementary materials, part 1.

### *2.2 Power calculations*

A simulation analysis was performed to assess the power of our study to detect a range of potential effect sizes for the risk of hospitalisation associated with the Delta variant of SARS-CoV-2, compared to the Alpha variant.

For this analysis, we follow a similar approach as described in [3]. In brief, the simulated datasets consisted of a binary response variable (Y) representing hospitalisation status and six predictor variables (X), each of them representing the covariate variables included in our final model (variant of SARS-CoV-2, age, sex, country of birth, underlying comorbidities and vaccination status). For simplification, variables “country of birth”, “underlying comorbidities” and “vaccination status” were simulated as binary variables, and “age” as a continuous variable. The input values used to simulate each of the predictor variables were selected to ensure that the simulated data resemble the distribution of our observed data set (Table S4). The binary response variable (Y) was generated from the Bernoulli distribution with probability of occurrence:

$\text{Prob}\{Y=1|X\}=1/(1+\exp(-X\beta))$

where $X\beta=X1\beta+\cdots+X6\beta6$

To evaluate what size effect of Delta variant that could be identified with sufficient power, the simulations were run for a range different values of coefficient βvariant. All the other regression coefficients (for age, sex, country of birth, risk and vaccination status) were set to the values obtained in our final model (Table S4). The simulation process was performed with R=1000. For each simulated dataset, we ran multiple logistic regression. The power (P) was estimated as the proportion of the results that detected the true value of βvariant, using a two-sided significance level $\alpha = 0.05$.

As presented in Table S5, with a sample size of 20,000 cases, a proportion of Delta variant of at least 40% and outcome proportion of 1.8%, we could detect a significant effect of 0.34 (odds ratio 1.4) with a statistical power of 86%.

**Table S4. Parameters' input and regression coefficients used in power calculation analysis**

| Predictor Variables | Distribution | Parameter | Input values | Coefficient* |
|---|---|---|---|---|
| Variant of SARS-CoV-2 (Delta vs Alpha) | Binomial | Probability(x=Delta variant) | 0.40 | 1.1-1.6 |
| Age (years) | Truncated Normal | Mean (SD)<br>Minimum–maximum | 31 (18)<br>0–104 | 0.76 |
| Sex | Binomial | Probability(x=male) | 0.53 | 0.16 |
| Risk Factors | Binomial | Probability(x=risk) | 0.1 | 0.65 |
| Vaccination status | Binomial | Probability(x=vaccinated) | 0.14 | -1.9 |
| Country of Birth | Binomial | Probability(x=Born in Norway) | 0.73 | 0.47 |

*Coefficient (β) of the predictors (X) with respect to the binary response variable (Y).

**Table S5. Statistical power to detect different effect sizes for the risk of hospitalisation associated with the Delta variant of SARS-CoV-2, compared to the Alpha variant.**

| Sample size | Proportion of Delta cases | Regression coefficient | Odds ratio | Power |
|---|---|---|---|---|
| 20 000 | 40% | 0.1 | 1.1 | 0.15 |
| | | 0.18 | 1.2 | 0.35 |
| | | 0.26 | 1.3 | 0.67 |
| | | 0.34 | 1.4 | 0.86 |
| | | 0.41 | 1.5 | 0.95 |
| | | 0.46 | 1.6 | 0.99 |

#### *2.3 Characteristics of cases with data on virus variant compared to all notified cases*

We assessed the representativeness of the cases with data on virus variant by comparing the characteristics of cases with data on virus variant and notified cases. The inclusion criteria are the same as for our main analysis. From all notified cases in Norway in the study period, we excluded 644 who did not have a national identity number, as well as 111 hospitalised with other main cause of admission than COVID-19 and three with unknown main cause of hospital admission. From all cases with data on virus variant, 92% (n=20,055) were included in our study cohort.

We found differences between cases with data on virus variant with regards to age group, county of residence, sampling week, and hospitalisation (Table S6). Differences in county and sampling week reflect the evolution of the outbreak as well as the availability and use of relevant PCR screening methodology for different virus variants at primary diagnostic laboratories. Differences in sampling week are also influenced by a delay in analysing samples from recent weeks, and variant data on more cases from recent weeks will be available in time. The proportion of cases with data on virus variant among hospitalised cases was slightly higher than among those not hospitalised (78% vs 73%) and very minor differences were observed among age groups and country of birth (affected by the “unknown” category). The above differences were considered minor for our study aim.

**Table S6: Characteristics of notified SARS-CoV-2 cases and cases with data on virus variant, Norway, 3 May – 15 August 2021**

| Characteristics | | All notified cases (% by characteristic) | Cases with data on virus variant | |
|---|---|---|---|---|
| | | | n | % of all notified |
| Total | | 29,628 (100%) | 21,691 | 73% |
| Sex | Female | 13,957 (47%) | 10,262 | 74% |
| | Male | 15,670 (53%) | 11,429 | 73% |
| | Unknown | 1 (0.01%) | 0 | 0% |
| | | | | P=0.132 |
| Age group | 0-24 years | 15,017 (51%) | 10,814 | 72% |
| | 25-44 years | 9,425 (32%) | 7,052 | 75% |
| | 45-64 years | 4,536 (15%) | 3,343 | 74% |
| | ≥65 years | 650 (2.2%) | 482 | 74% |
| | | | | P<0.0001 |
| Norwegian born | Yes | 20,866 (70%) | 15,206 | 73% |
| | No | 8,527 (29%) | 6,321 | 74% |
| | Unknown | 235 (0.8%) | 164 | 70% |
| | | | | P=0.043 |
| Risk for severe COVID-19 * | No underlying comorbidities | 27,009 (91%) | 19,758 | 73% |
| | Medium risk comorbidity | 2,435 (8.2%) | 1,795 | 74% |
| | High risk comorbidity | 184 (0.6%) | 138 | 75% |
| | | | | P=0.718 |
| Vaccine status at date of positive test | Not vaccinated | 22,496 (76%) | 16,490 | 73% |
| | Vaccinated with 1 dose <21 days before positive test | 2,514 (8.5%) | 1,814 | 72% |
| | Partially vaccinated | 3,412 (12%) | 2,526 | 74% |
| | Fully vaccinated | 1,206 (4.1%) | 861 | 71% |
| | | | | P=0.193 |
| Period of diagnosis | Weeks 18-20 | 8,601 (29%) | 6,513 | 76% |
| | Weeks 21-23 | 5,182 (17%) | 3,897 | 75% |
| | Weeks 24-26 | 3,659 (12%) | 2,638 | 72% |
| | Weeks 27-29 | 3,597 (12%) | 2,689 | 75% |
| | Weeks 30-32 | 8,589 (30%) | 5,954 | 69% |
| | | | | P<0.0001 |

| | | | | |
|---|---|---|---|---|
| County of residence | Agder | 2,467 (8.3%) | 2,251 | 91% |
| | Innlandet | 1,620 (5.5%) | 1,081 | 67% |
| | Møre and Romsdal | 875 (3.0%) | 474 | 54% |
| | Nordland | 547 (1.9%) | 224 | 41% |
| | Oslo | 5,614 (19%) | 4,540 | 81% |
| | Rogaland | 2,447 (8.3%) | 1,770 | 72% |
| | Troms and Finnmark | 1,313 (4.4%) | 577 | 44% |
| | Trøndelag | 1,948 (6.6%) | 1,379 | 71% |
| | Vestfold and Telemark | 3,025 (10.2%) | 2,258 | 75% |
| | Vestland | 3,217 (10.9%) | 2,418 | 75% |
| | Viken | 6,543 (22%) | 4,712 | 72% |
| | Unknown | 12 (0.04%) | 7 | 58% |
| | | | | P<0.0001 |
| Hospitalised with COVID-19 as main cause of hospitalisation | Yes | 475 (1.6%) | 369 | 78% |
| | No | 29,153 (98%) | 21,322 | 73% |
| | | | | P=0.026 |

Note: p-values presented are from chi-square tests. * Risk for severe disease based on underlying comorbidities that are associated with a moderate or high risk of serious illness regardless of age. Details on the definitions of medium and high risk categories are provided in supplementary materials, part 1.

*2.4 Distribution of vaccine type in the study cohort*

**Table S7: Distribution of vaccine type among vaccinated SARS-CoV-2 cases in the study cohort, by virus variant, Norway, 3 May – 15 August 2021**

| | Alpha cases | | | | Delta cases | | | |
|---|---|---|---|---|---|---|---|---|
| | Partially vaccinated | | Fully vaccinated | | Partially vaccinated | | Fully vaccinated | |
| Vaccine type | N | % | N | % | N | % | N | % |
| Comirnaty | 304 | 66 | 153 | 86 | 1,584 | 82 | 449 | 71 |
| Spikevax | 43 | 9.3 | 3 | 1.7 | 303 | 15 | 67 | 11 |
| Vaxzevria | 111 | 24 | 0 | 0 | 9 | 0.5 | 5 | 0.8 |
| Vaxzevria + Comirnaty/ Spikevax | - | - | 19 | 11 | - | - | 96 | 15 |
| Janssen | 0 | 0 | 3 | 1.7 | 0 | 0 | 12 | 1.9 |
| Unknown | 3 | 0.7 | 0 | 0 | 36 | 1.9 | 0 | 0 |
| Totalt | 461 | 100% | 178 | 100% | 1,925 | 100% | 629 | 100% |

*2.5 Stratified estimates for the risk of hospitalisation by vaccination status*

In order to investigate further the association between vaccination status and the risk of hospitalisation, we conducted a stratified analysis for some of the variables in our main model (age, underlying comorbidities and variant) (Table S8). This also allows us to investigate the interaction detected between age group and vaccination status.

In the age group 0-24 years, no difference in the risk of hospitalisation among partially and fully vaccinated cases was observed, compared to unvaccinated cases, although this needs to be interpreted in light of our small sample size and rarity of the outcome for both these groups, with 754 partially vaccinated cases (of which three hospitalised) and 90 fully vaccinated cases (of which one hospitalised) in this age group. Among 8,580 unvaccinated cases aged 0-24 years in our dataset, 20 were hospitalised. The same can be said of fully vaccinated cases aged 25-44 years (234 cases, of which one hospitalised) and partially vaccinated persons with a high risk comorbidity (25 cases, of which three hospitalised).

**Table S8. Stratified estimates using multivariable binomial regression for the risk of hospitalisation by vaccination status among SARS-CoV-2 cases, Norway, 3 May – 15 August 2021**

| | | Adjusted risk ratio (95%CI) for hospitalisation | | | |
|---|---|---|---|---|---|
| | | Not vaccinated | Vaccinated with 1 dose <21 days before positive test | Partially vaccinated | Fully vaccinated |
| Overall | | Ref | 0.78 (0.56-1.08) | 0.28 (0.18-0.41) | 0.24 (0.15-0.39) |
| *Stratified by:* | | | | | |
| Age group | 0-24 years | Ref | * | 1.15 (0.32-4.17) | 3.21 (0.42-24.9) |
| | 25-44 years | Ref | 0.57 (0.27-1.18) | 0.16 (0.05-0.51) | 0.17 (0.02-1.23) |
| | 45-64 years | Ref | 1.16 (0.78-1.75) | 0.16 (0.08-0.34) | 0.29 (0.12-0.70) |
| | >65 years | Ref | 0.30 (0.14-0.65) | 0.30 (0.17-0.53) | 0.18 (0.10-0.32) |
| Risk for severe COVID-19 ** | No underlying comorbidities | Ref | 0.71 (0.47-1.07) | 0.29 (0.18-0.48) | 0.21 (0.09-0.46) |
| | Medium risk comorbidity | Ref | 0.84 (0.48-1.48) | 0.15 (0.06-0.40) | 0.29 (0.14-0.60) |
| | High risk comorbidity | Ref | 0.92 (0.26-3.31) | 0.40 (0.12-1.32) | 0.18 (0.06-0.50) |
| Variant | Delta | Ref | 0.36 (0.17-0.80) | 0.23 (0.13-0.42) | 0.21 (0.11-0.41) |
| | Alpha | Ref | 0.92 (0.64-1.32) | 0.28 (0.15-0.50) | 0.30 (0.15-0.61) |

* There were no hospital admissions among 562 cases aged 0-24 years who were vaccinated with 1 dose <21 days before positive test. ** Risk for severe disease based on underlying comorbidities that are associated with a moderate or high risk of serious illness regardless of age. Details on the definitions of medium and high risk categories are provided in supplementary materials, part 1.

*References*

1. Sheikh A, McMenamin J, Taylor B, Robertson C. SARS-CoV-2 Delta VOC in Scotland: demographics, risk of hospital admission, and vaccine effectiveness. Lancet. 2021. doi: 10.1016/S0140-6736(21)01358-1.
2. Twohig KA, Nyberg T, Zaidi A, Thelwall S, Sinnathamby MA, Aliabadi S, et al. Hospital admission and emergency care attendance risk for SARS-CoV-2 delta (B.1.617.2) compared with alpha (B.1.1.7) variants of concern: a cohort study. Lancet Infect Dis. 2021. https://doi.org/10.1016/S1473-3099(21)00475-8
3. Gastanaga VM, McLaren CE, Delfino RJ. Power calculations for generalized linear models in observational longitudinal studies: a simulation approach in SAS. Comput Methods Programs Biomed. 2006;84(1):27-33.
